# Rationale of acupuncture for stabilizing blood pressure fluctuation during total laparoscopic hysterectomy (ASBP): a parallel grouped, randomized clinical trial

**DOI:** 10.1101/2024.07.25.24310977

**Authors:** Joohyun Lee, Ju-Won Roh, Kyung-Hee Han, Min-Jeong Kim, Young Jeong Na, Bo Seong Yun, Jee Young Lee

## Abstract

**Introduction:** Reducing blood pressure fluctuations during surgery is a significant goal for anesthesiologists. Acupuncture may be a non-invasive intervention to reduce blood pressure fluctuations but has not yet been studied. This study aims to determine whether acupuncture can be used to reduce blood pressure fluctuations during surgery, especially in the early stages.

**Methods and analysis:** This is a prospective, single-center, randomized controlled clinical trial with a parallel-group design. Thirty adult patients scheduled to undergo total laparoscopic hysterectomy are eligible. Participants who consent will be randomly assigned in a 1:1 ratio to the acupuncture or placebo group. They will be followed up for at least 14 days to assess the safety of the intervention, general anesthesia, and surgery. We will compare the differences between the highest and lowest mean blood pressures from anesthesia induction to the post-incision period as the primary endpoint. As secondary outcomes, systolic, diastolic, and mean blood pressures will be compared at each pre-determined time point. Incidence of hypotension, hypertension, tachycardia, and bradycardia will be counted separately. The use of remifentanil at the early stage of surgery, the rate of surgical discontinuation, and the length of hospital stay will be assessed as surrogate indicators of stable general anesthesia and surgical procedures. For patient-reported outcomes, Spielberger’s State-Trait Anxiety Inventory and EuroQoL-5 Dimensions–5 Levels will evaluate the change in anxiety and overall quality of life. Another non-pharmacological intervention may contribute to surgery by maintaining blood pressure within a stable range during the early postoperative period.

**Ethics and dissemination:** The study will be conducted in accordance with the Declaration of Helsinki, and approved by the Institutional Review Board of CHA Ilsan Medical Center (ICHA 2022-11-010, date of approval 2023-01-03). This study was registered at Clinicaltrials.gov (registration identifier: NCT05720884) and CRiS (registration identifier: KCT0009149). The publication is scheduled for December 2025. Data deposition is scheduled to occur.

**Strengths and limitations of this study:**

1. Reducing blood pressure fluctuation is important during surgical process.
2. Considering pharmacological interactions, non-pharmacological interventions are preferable for managing blood pressure fluctuation.
3. Acupuncture has some potential for stabilizing blood pressure.

## INTRODUCTION

Various factors, such as the use of agents that induce anesthesia, surgery-induced stress, or hemorrhage, contribute to blood pressure fluctuations during surgery. Since blood pressure lower or higher than normal range can affect patient prognosis, reducing blood pressure fluctuations during surgery is an important goal for anesthesiologists [1,2]. A previous study reported that both hypotension and hypertension increased the oxygen demand in peripheral organs, postoperative complications, and mortality [3].

Blood pressure fluctuations occurring during preoperative management, which covers anesthesia induction for tracheal intubation and skin incision and the early stages of surgery, develop rapidly in a very short period. Thus, it is highly important to expect a given stimulation and its intensity beforehand and control the pain level [4]. This procedure requires skilled anesthesiologists with full experience in the related work. Anesthesiologists should perform close patient evaluations before surgery and provide preoperative management to prevent rapid blood pressure fluctuations during surgery. Various drugs such as premedication with benzodiazepines, midazolam, alpha-2 adrenergic agonists, or ketamine were included to review the most appropriate drugs [5–7].

However, premedication in adult surgical patients should be used cautiously, considering its risks and benefits [8]. Premedication can cause adverse reactions such as over-sedation and dizziness [9]. As oversedation threatens patient safety by causing hypoxia, close observation is required from the time of sedatives administration to entering the operating room, and this can act as throughput.

It is favorable that some non-pharmacological interventions, rather than pharmacological ones, can contribute to hemodynamic stability in patients undergoing surgery, due to their minimized interactions with the drugs used during the surgical procedure. Acupuncture could be a candidate for these non-phamacological interventions. Therefore, this prospective clinical study is designed to evaluate the efficacy of acupuncture on blood pressure fluctuations during surgery in adult patients, especially from the induction time to the post-incision moment, by comparing it with a placebo control group.

## METHODS AND ANALYSIS

### Study Design

This is a prospective, single-center, randomized controlled clinical trial with a parallel-group design for blood pressure fluctuations during general anesthesia. Thirty adult patients scheduled to undergo a total laparoscopic hysterectomy will be included in this study. Participants who voluntarily consent to participate in the clinical trial will be randomly assigned in a 1:1 ratio to the acupuncture or placebo group. Eligible participants will be followed up for at least 14 days to assess the safety of the intervention, general anesthesia, and surgery.

The study protocol adheres to both the Standard Protocol Items: Recommendations for Interventional Trials (SPIRIT) guidelines and checklist and Standards for Reporting Interventions in Clinical Trials of Acupuncture (STRICTA) guideline (Supplement 1). All study procedures will be performed in accordance with the Declaration of Helsinki and Korean Good Clinical Practice Guidelines. The study protocol has been approved by the Institutional Review Board (ICHA 2022-11-010) and registered and updated at Clinicaltrials.gov (NCT05720884), the Clinical Research Information Service, and the South Korean registration service for clinical trials (KCT0009149). This article describes the study based on protocol version 1.1, dated on 2023-01-14; the latest protocol will be updated in a timely manner on both registration sites. Study flow diagram is presented in Figure 1.

**Figure 1.**
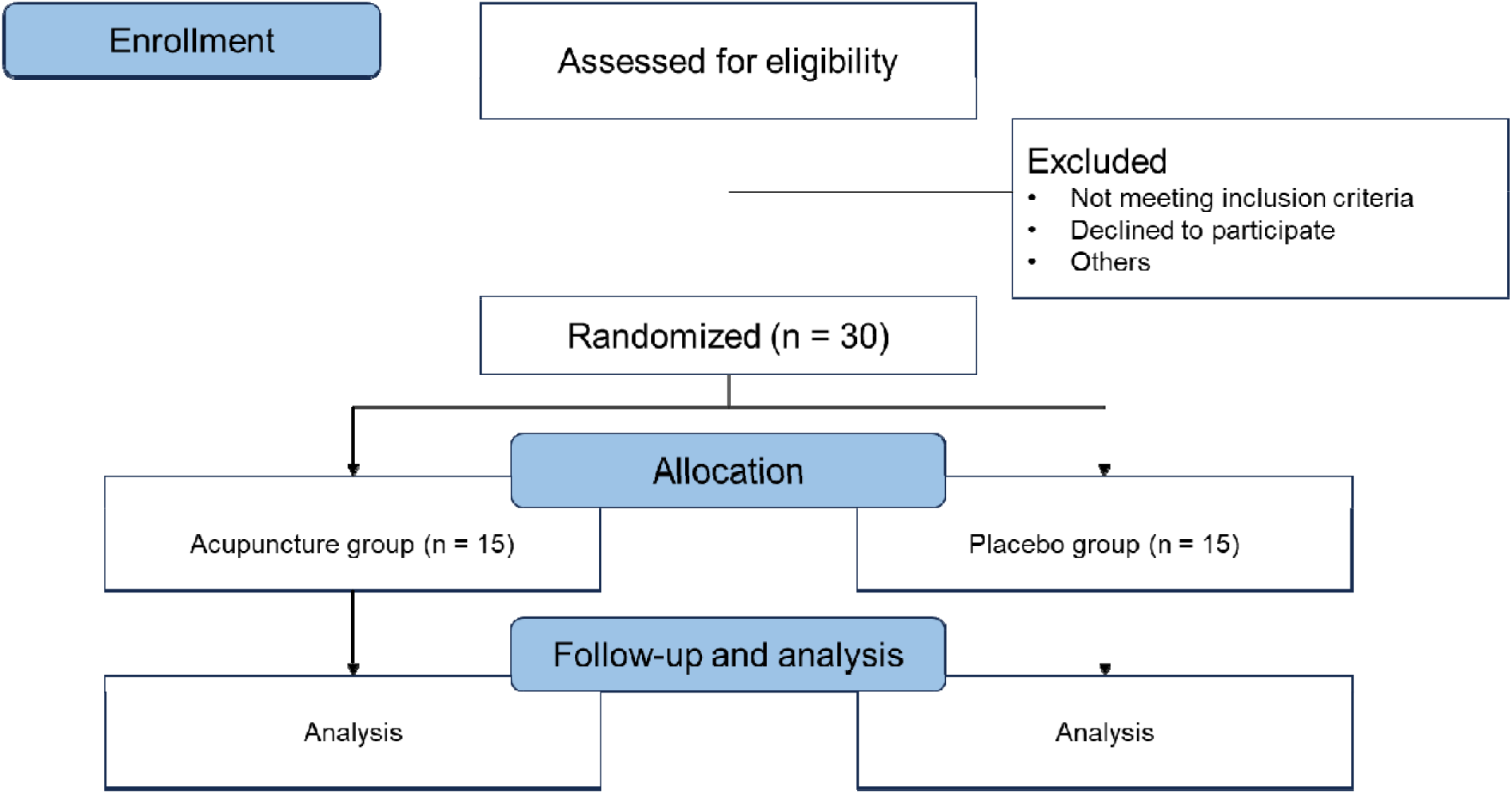
Flow diagram of the trial

### Eligibility

Participants scheduled to undergo surgical removal of the uterus for uterine fibroids and had the expected date of total laparoscopic hysterectomy by a gynecologist are eligible. The specific inclusion criteria are as follows: 1) age between 19 and 69 years, 2) American Society of Anesthesiologists (ASA) class I or II, 3) requirement for surgical removal of the uterus by a gynecology specialist, and 4) scheduled to undergo total laparoscopic hysterectomy in advance. All participants will be required to understand and agree to the study protocol. Written informed consent will be obtained from all participants by a delegated researcher or research coordinator.

Participants under following conditions or situations are ineligible: 1) expected survival within 3 months; 2) emergency operation which was not scheduled in advance; 3) hypertension or hypotension that can significantly interfere with the study result; 4) diagnosis of arrhythmia, such as atrial fibrillation, frequent ventricular or supraventricular premature beats; 5) diagnosis of heart failure or valvular disease; 6) anemia of hemoglobin <12 g/dL; 7) incapable of surgery due to hemodynamic or medical reasons other than stated above; 8) incapable of receiving acupuncture treatment on determined location; 9) current use of gonadotropin-releasing hormone receptor agonists; 10) current use of drugs that may interfere with the result, including steroids, immunosuppressants, and psychiatric disorders; 11) significant comorbidities that may interfere with the interpretation of intervention efficacy or results, such as stroke, myocardial infarction, kidney disease, dementia, or epilepsy; 12) pregnant, planning to be pregnant within study period, or breastfeeding; 13) previous participation in any other clinical trial within 1 month of participation, planning to participate another clinical trial within 6 months after enrollment date, or planning to participate another clinical trial on follow-up period; 14) fail to write the informed consent form voluntarily; and 15) being deemed to be unsuitable for participation.

### Randomization and Allocation

Eligible participants who sign the informed consent form will be randomly assigned to the acupuncture or placebo group in a 1:1 ratio. Random sequences will be generated by permuted block randomization of block size 2 or 4 using the random allocation tool in R software 4.1 (R Foundation for Statistical Computing, Auckland, New Zealand). Random sequence generation will be performed and random sequences will be determined by an independent statistician prior to enrollment. Only the statistician who generated the random sequence will be aware of the full randomization sequence.

Each random sequence will be sealed in an opaque envelope and stored in a double-locked cabinet. The delegated researcher or research coordinator will explain the trial process to candidate, and get written informed consent, and open the randomization envelope for each participant and assign them to an appropriate group.

Intervention researchers will perform acupuncture or placebo acupuncture according to their assigned group. The intervention researcher will not be blinded because the researcher should perform different interventions according to the group. However, we will separate the person and location to maintain blindness as robustly as possible. Both the participant and the assessor will be blinded, and the location where the intervention is performed will be separate from that where the assessment will be performed. Interventions will be performed in the ward, and assessments will be performed in the operating room.

### Intervention

Acupuncture with a disposable, sterilized, stainless steel needle of 0.20 mm × 30 mm size (Dongbang Medical, Boryeong, Republic of Korea) will be used for both groups. Ten acupoints (PC6, PC5, SP3, KI3, and ST36) will be chosen bilaterally for the intervention group, also known as the acupuncture group. The depth of acupuncture will be varied among the participants. After acupuncture needles are inserted, they will be manually manipulated to obtain a pre-determined sensation, the De Qi. The acupuncture will remain inserted for 20 min. Ten other points that are not official acupoints and are not on the median nerve or peroneal nerve will be chosen for the placebo group. Acupuncture or placebo will be administered 2 times during the first and second visits. Acupuncture or placebo acupuncture will be administered by a qualified acupuncturist, who is a physician in Korean medicine with 10 years of clinical experience.

### Study Process

After a sufficient explanation of the purpose and methods of this clinical study by a researcher, subjects who voluntarily decide to participate and sign a written consent form will be evaluated for eligibility at the screening visit. Eligible subjects will be randomly assigned to the acupuncture and placebo control groups according to the randomization. Acupuncture treatment will be performed twice, one day before surgery (V1) and on the day of the surgery (V2).

All participants will fast for 6–8 h prior to surgery. After entering the operating room (V3), electrocardiography, pulse oximetry, and monitoring of the partial pressure of end-tidal carbon dioxide (PETCO2) (IntelliVue MX550, Phillips North America Corporation, Andover, MA, USA) will be continuously performed. Automated oscillometric measurements of blood pressure at 2.5-min intervals and Train of Four (TOF) (ToFscan®, Drager Technologies, Canada) will be also measured. Throughout the surgery, these data will be downloaded to personal computers using RS232C cables.

After checking the initial vital signs (heart rate, blood pressure, and oxygen saturation) (P1), anesthesia will be induced intravenously with propofol 1–2.5 mg/kg and remifentanyl (0.01-0.1 ng/kg/min). Mask ventilation (P2) with desflurane will be performed after the administration of rocuronium (0.6–1 mg/kg). Once the TOF ratio reaches <2, the anesthesiologist will perform tracheal intubation (P3). The lungs of the subjects will then be ventilated with desflurane and oxygen in air (FiO2=0.5), and the tidal volume and ventilation rate will be adjusted to maintain PETCO2 between 35 and 45 mmHg. Then, an additional venous tube will be secured (P4).

With the patient in the lithotomy position and under general anesthesia, the abdomen will be prepared, painted, and draped in the usual manner (P5). The uterine elevator will be inserted. A skin incision will be made (P6), and a trocar will be inserted. CO2 gas will be infused to create a pneumoperitoneum at a pressure of 12 mmHg (P7). After entering the abdominal cavity, the patient’s position will be changed to Trendelenburg. The pelvic cavity and entire abdomen will then be examined. Bilateral pelvic sidewall triangles will be opened parallel to the infundibulopelvic ligament and ureteral dissection will be performed. The ureteral course will be identified, and the right round and ovarian ligaments will be coagulated with a bipolar endocoagulator and cut with endoscissors. The left uterine ligaments will be manipulated using the method described above. The bladder flap will be pushed and the posterior broad ligament will be mobilized. The bilateral uterine vessels will be skeletonized, coagulated, and cut. Circumferential colpotomy will be performed using unipolar scissors, the uterus will be removed transvaginally, and the vaginal cuff will be closed using continuous intracorporeal sutures. Hemostasis and ureteral peristalsis will be ensured, and the abdominal cavity will be irrigated with normal saline. The subcutaneous tissue and skin will be closed layer-by-layer.

During the procedure, the infusion rate of remifentanil and the concentration of desflurane (6.0–7.0 vol%) will be adjusted to maintain stable hemodynamic values (systolic blood pressure >80 mmHg; heart rate >45 beats/min), and an appropriate depth of anesthesia (>1 minimal alveolar concentration). If necessary, ephedrine or atropine will be administered to maintain stable hemodynamic values. All decisions will be made by a qualified anesthesiologist with at least 10 years of clinical experience.

The neuromuscular blockade will be reversed by administering sugammadex 2–5 mg/kg at the end of the surgery, and the patient will be confirmed to be fully awake (TOF>0.9) and transferred to the post-anesthetic care unit (P8).

By participating in this clinical study, the study participants will not be restricted from receiving rescue medicine required in the surgical process. If rescue medicine or rescue treatment is administered, the researcher will record the combination therapy and rescue medicine in the case report form.

Since the intervention was processed over a short term, strategies for study adherence were deemed unnecessary. However, to improve adherence, the follow-up survey was made available both online and offline. The online platform data is only accessible to the principal Investigator and the delegated research coordinator. Other data will be electronically locked or stored in a document archive accessible only to researchers for the duration of the study and for three years after its completion.

### Outcome Measures

To investigate the efficacy of acupuncture on initial blood pressure fluctuations during laparoscopic hysterectomy, we will compare the difference between the highest and lowest mean blood pressures from the induction of anesthesia to the post-incision period as the primary endpoint. As secondary outcomes, systolic, diastolic, and mean blood pressures will be compared at each pre-determined time point. The incidences of hypotension (systolic blood pressure <90 mm Hg or 80% of the baseline), hypertension (systolic blood pressure >160 mm Hg or 120% of the baseline), tachycardia, and bradycardia will be recorded. The total use of remifentanil, surgical discontinuation, and length of hospital stay will be assessed as surrogate indicators of stable general anesthesia and surgical procedures.

For patient-reported outcomes, Spielberger’s State-Trait Anxiety Inventory (STAI) and EuroQoL-5 Dimensions – 5 Levels (EQ-5D-5L) will be used to evaluate changes in anxiety and overall quality of life. The STAI was first validated in 1996 [10,11]. The STAI consists of 40 questions divided into two categories: 20 items assessing state anxiety and 20 items assessing trait anxiety. State anxiety (SAI) refers to experiencing anxiety in the moment, and trait anxiety (TAI) refers to the type and characteristics of anxiety.

The EQ-5D-5L will be used to assess the effects on the patients’ quality of life. The questionnaire consists of questions in five areas: mobility, self-care, usual activities, pain, and anxiety/depression. The EQ-5D-5L assesses the patient’s current state of health. Responses are provided on a 5-point Likert scale [12].

Study process and time points are presented in Table 1.

**Table 1.**
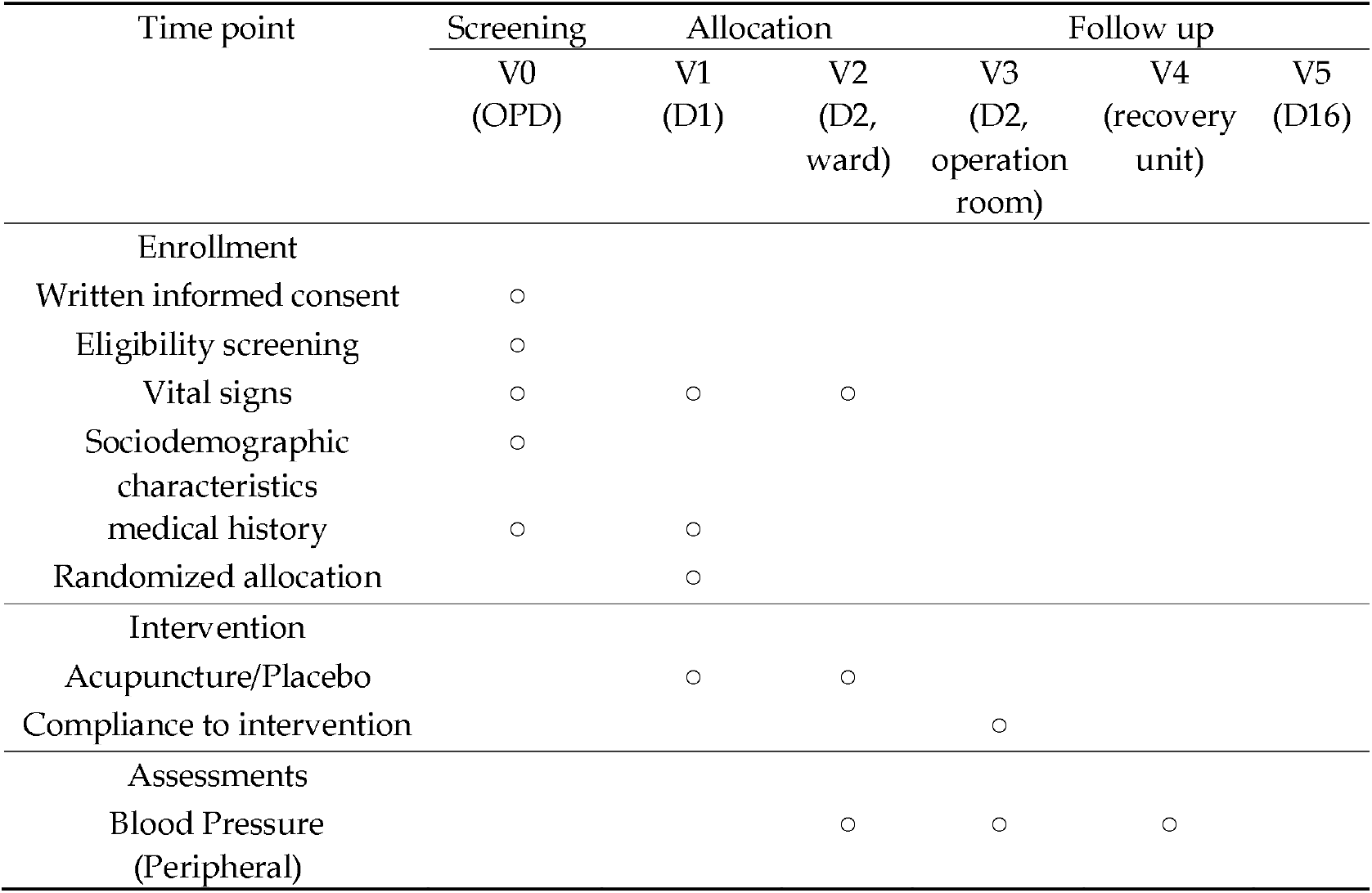
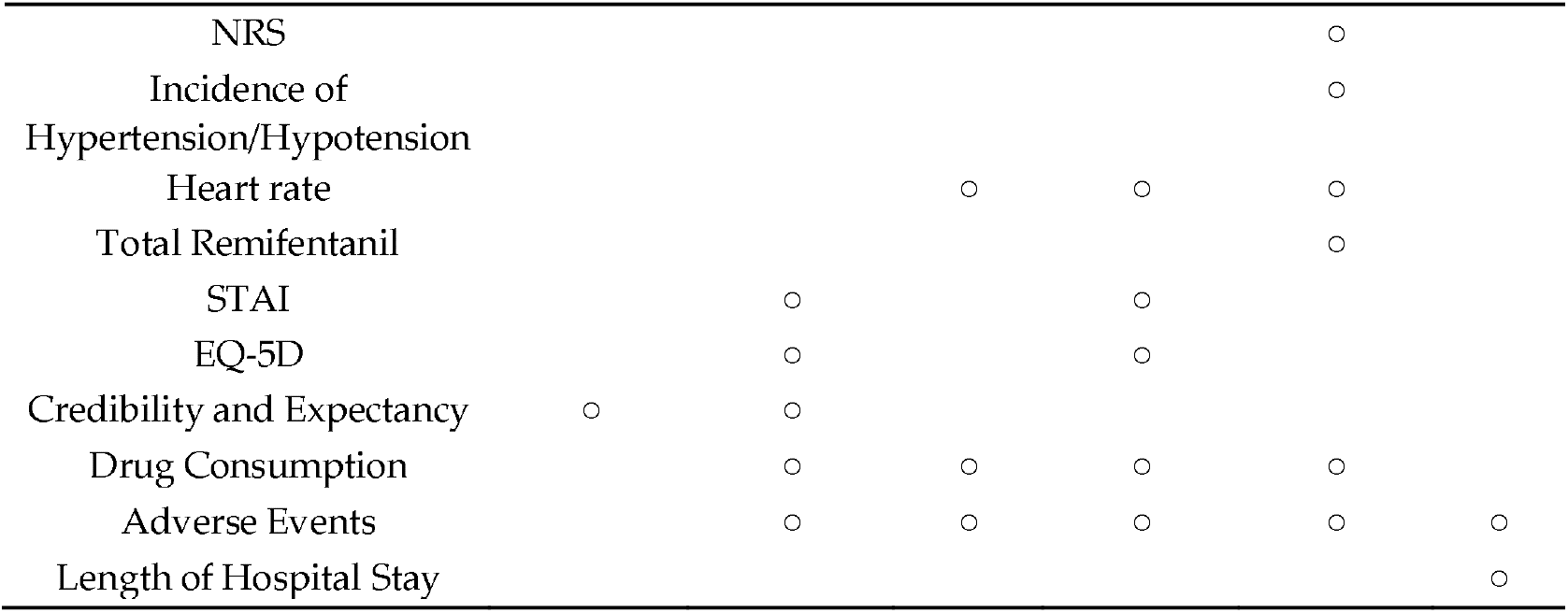
Timetable and outcome measurements.

### Concomitant care

Participants are not restricted from concomitant care, similar to conditions in an actual clinical environment. They can choose any type of combination therapy and any necessary salvage treatment throughout the entire hospitalization period. Researchers will document those therapies in the case report form. The documentation of these therapies will include the type, dosage, route of administration, frequency, and purpose.

### Safety

Safety will be evaluated by grading the severity of symptoms or medical conditions. Adverse events will be defined according to MedDRA 24.0, and causality will be assessed according to the World Health Organization-Uppsala Monitoring Centre (WHO-UMC) Causality Assessment. The severity of subjective and objective symptoms will be evaluated according to the Common Terminology Criteria of Adverse Events version 6.0.

The clinical study is covered by liability insurance to compensate for any foreseeable or unforeseeable harm that may occur during the research process.

### Sample Size

No previous studies have compared the efficacy of acupuncture treatment for target conditions and blood pressure fluctuations during general anesthesia in total laparoscopic hysterectomy. This is a pilot study to evaluate the feasibility of a follow-up study, setting an allocation ratio of 1:1 as the minimum unit of 12 patients deemed necessary for the pilot study. Considering a dropout rate of 20%, total 30 participants will be recruited.

### Data and Safety Monitoring Board

This clinical study involves a data and safety monitoring board (DSMB) consisting of at least one medical clinician and a statistician. The composition and operation of the DSMB will be in accordance with the standard operating procedure of the trial. Study monitoring is scheduled to be conducted once a year. If protocol modification was recommended, appropriate amendment or response will be provided. If a suspected unexpected serious adverse reaction case occurs or the DSMB suspects a significant harmful association with the intervention and requests unblinding, the intervention researcher will unblind the allocation to other researchers and the committee, and submit the unblinded interim results.

### Statistical Analysis

Both intention-to-treat (ITT) and per-protocol (PP) analyses will be performed, with ITT as the primary analysis. For the PP analysis, the study participants who successfully completed the surgery will be analyzed separately. Missing values will be processed through a Mixed Model for Repeated Measures in the case of a Linear Mixed Model, and multiple imputations will be performed to obtain estimates and standard errors by setting 10 imputation sets for additional analysis.

Sociodemographic characteristics and treatment expectations of the study participants will be evaluated for each group. Continuous variables will be expressed as mean (standard deviation) or median (quartile), and the differences between the two groups will be compared using the Student’s t-test or Wilcoxon rank-sum test according to the distribution. Categorical variables will be expressed as frequencies (%), and a chi-square test or Fisher’s exact test will be performed.

The main endpoint of this clinical trial is the difference between the highest and lowest mean blood pressures measured during the period from anesthesia induction to the post-incision period. The Student’s t-test will be used to test whether the difference between the two groups in the main endpoint is statistically significant. Differences between the two groups will be compared based on the Student’s t-test or Wilcoxon rank-sum test, unless otherwise specified, for hemodynamic stabilization time, incidence of hypotension and hypertension, comparison of drug use, and other efficacy endpoints, including STAI. The two groups will be compared by estimating the ratio between the blood pressure measured at each time point and that measured at V2, using the blood pressure at V2 as the baseline. Since blood pressure is measured repeatedly, it is not possible to assume independence between measurements; thus, generalized estimating equations will be used. To estimate this ratio, the measurements will be log-transformed and included in the model. The two groups will be compared using the same method for the pulse rate measured at V2 and the pulse rate at each time point. To quantify the length of hospitalization in both groups, we will use Kaplan-Meier Estimation to estimate the time until 50% of the participants are discharged. The log-rank test will be used to determine differences in the distribution of discharge times between the two groups.

The significance level of all analyses is set at 0.05. All statistical analyses will be performed using SAS 9.4 (© #SAS Institute, Inc., Cary, NC, USA) or R software 4.1 (R Foundation for Statistical Computing, Auckland, New Zealand).

## DISCUSSION

The primary role of an anesthesiologist during surgery extends beyond maintaining unconsciousness in patients to actively minimizing physical stress induced by noxious stimuli. These stimuli include not only the surgical stimulation itself but also the medications administered to maintain unconsciousness or alleviate surgical pain. While essential for their intended purposes, both anesthetics and analgesics disrupt the body’s homeostasis necessary for life and can therefore be classified as injurious stimuli. Administration of these medications commonly leads to intraoperative hypotension, which can be exacerbated by changes in patient positioning or surgical hemorrhage. Even in healthy individuals without underlying diseases, intraoperative hypotension adversely affects postoperative outcomes [13,14]. Hence, it is advised to avoid prolonged episodes of hypotension exceeding 10 min during non-cardiac surgeries [15].

During surgery, the increase in blood pressure is a sign of the stress response triggered by noxious stimuli in the patient’s body. This response may indicate inadequate dosing of analgesics administered during surgery. Failure to effectively manage pain-induced stress can result in increased postoperative analgesic requirements, leading to potential complications such as respiratory depression, nausea, vomiting, and prolonged hospital stays [16]. Additionally, intraoperative hypertension not only indicates inappropriate pain management but also increases the risk of major adverse cardiovascular events (MACE) itself [3], making it a clinical situation requiring proactive management by an anesthesiologist similar to intraoperative hypotension.

To avoid hypotensive and hypertensive episodes during surgery, anesthesiologists vigilantly monitor both the patient’s hemodynamics and surgical progress throughout the anesthesia, titrating anesthetic and analgesic dosages accordingly. Real-time fluctuations in intraoperative blood pressure depend on the progress of surgery and drug kinetics. It is known that maintaining intraoperative blood pressure within 10% of preoperative levels significantly reduces the risk of post-operative organ dysfunction [17,18]. Therefore, anesthesiologists must actively manage medications and interventions affecting blood pressure, not only intraoperatively but also in the preoperative phase.

Since acupuncture is known to be effective in dilating blood vessels and improving circulation in peripheral blood flow, studies have been conducted to investigate whether acupuncture therapy affects blood flow and pressure [19]. According to previous studies, acupuncture acts on the cerebrum, which regulates blood pressure, as shown by functional magnetic resonance imaging, and is known to be effective in regulating essential hypertension [20]. However, the lowering effect of acupuncture therapy on blood pressure is not clinically or directly strong in practice. Studies by Flachskampf et al. and Yin et al. used acupuncture therapy for 42 and 56 days, respectively, and significant effects were observed in the acupuncture group compared to the sham acupuncture control group [21,22]. However, no significant differences were observed in studies that used acupuncture therapy for 15 or 25 days [23,24]. Overall, although the long-term use of acupuncture therapy has blood pressure-lowering effects of 4–10 mmHg in patients with uncontrolled hypertension, the effects of short-term acupuncture therapy on blood pressure fluctuations have not yet been discovered. Therefore, we aim to investigate whether acupuncture therapy stabilizes blood pressure fluctuations. More specifically, this prospective, exploratory, pilot study is designed to explore whether acupuncture therapy lowers blood pressure fluctuations in the early stages of surgery compared with a placebo group. In terms of safety, acupuncture therapy is considered a rather safe non-pharmacological, minimally invasive option. When using acupuncture therapy at the level to be used in this study, the incidence of adverse events was approximately 9.4% or less, half of which were local subcutaneous hemorrhages. Of the 1,000 patients with adverse events, 1.14 required treatment [25].

Several considerations are required to establish the outcome indicators that most accurately define the reduction of blood pressure fluctuations during surgery. The first consideration was whether to choose a relative or absolute value to maintain the blood pressure. In early 2000, if the goal was to maintain blood pressure level within 2–30% of the baseline while monitoring the relative value, it would be considered physiologically significant, especially in patients with chronic hypertension or elderly patients [26]. However, establishing the baseline for relative value presents challenges due to the lack of consensus on whether baseline blood pressure should be determined during preoperative evaluation or preinduction. Preinduction blood pressure, typically measured upon entering the operating room, differs from blood pressure recorded during the perioperative evaluation. Among 4,408 patients aged ≥60 undergoing noncardiac surgery, 61% exhibited a mean blood pressure that was 10 mmHg lower during preoperative evaluation compared to preinduction mean arterial pressure [27]. Although the blood pressure obtained during the preoperative evaluation is used as a reference value, normal blood pressure levels vary depending on individual conditions and the circadian cycle, resulting in different normal values in the same patient according to the preoperative evaluation time [28]. Therefore, absolute values are more commonly used, and the present study will also use absolute values to investigate the primary outcomes. In more specific figures, the underline of the mean arterial pressure was set at 65 mmHg, which was considered an indicator of organ perfusion, and the upperline was set at 80 mmHg, which is known to increase the incidence of MACEs [29].

Despite the anesthesiologist’s effort to maintain the target blood pressure level during surgery, each person responds differently to analgesics and stimuli from tracheal intubation and surgical stimulation, and situations during surgery continue to change. For patients expected to experience massive hemorrhage or underlying diseases that may increase cardiovascular morbidity, invasive monitoring by using A-line wave monitoring is required. Considering the ethical issues, an automated non-invasive blood pressure device (NIBP), which is a non-invasive monitoring device, can also be used. To select a patient group with fewer systemic problems or vascular issues and few sexual and physiological variations, a patient group with leiomyoma requiring laparoscopic hysterectomy has been selected. Based on a previous study recommending measurement of NIBP every 2.5 min for reducing the incidence of unrecognized data during endotracheal intubation, NIBP was set every 2.5 min to closely monitor rapidly changing blood pressure in the early stages of anesthesia and surgery [30].

Meanwhile, the initial period after anesthesia induction and the start of surgery is characterized by significant blood pressure fluctuations. Following loss of consciousness due to anesthetic drugs, there is a period of sympathetic withdrawal leading to a decrease in blood pressure, followed rapidly by a sharp increase due to nociceptive stimuli such as endotracheal intubation. During periods without stimulation after intubation, blood pressure generally decreases due to the effects of anesthetic agents administered for the maintenance of general anesthesia; however, the initial surgical stimuli such as skin incision promptly elevate blood pressure again. Therefore, the blood pressure fluctuations in the early stages of surgery can serve as a representative indicator of overall blood pressure changes during surgery. This study aims to carefully observe this period to assess the efficacy of nursing interventions in managing blood pressure fluctuations.

In particular, laparoscopic hysterectomy is a surgical approach that demonstrates early-stage blood pressure fluctuations. Following general anesthesia induction increases in blood pressure and heart rate commonly occur due to stimulation of the sympathetic nervous system during the processes of insufflation and positioning for surgical field visualization [31,32]. In contrast, the vasovagal reflex triggered by peritoneal insufflation may result in severe bradycardia, requiring a high level of anesthesia precautions [33].

During this period, anesthesiologists aim to maintain optimal blood pressure by carefully adjusting the administration of analgesics. The timing, rate of administration, and dosage of supplemental opioids should be tailored to the patient’s condition and the anticipated duration of the operation to avoid complications [34]. For example, to prepare for tracheal intubation and surgical nociceptive stimuli, the serum concentration of analgesics is adjusted beforehand to increase the antinociceptive level. Conversely, during periods predicted to be without stimulation or when the parasympathetic nervous system predominates, the concentration of analgesics is reduced [35]. Due to the time lag between the administration of analgesic and achieving the target concentration in the body, a preemptive action should be taken for nociceptive stimuli that are predicted. If the time points of the procedure are not well synchronized, significant variations in blood pressure due to the presence or absence of stimuli can occur, potentially impacting patient outcomes adversely. The opioid analgesic remifentanil, which will be utilized in our study, has a short half-life and, like other opioids, not only delays emergence from general anesthesia but can also lead to postoperative nausea and vomiting, as well as opioid-induced hyperalgesia in the recovery room [36].

Due to its short half-life, remifentanil is commonly administered via infusion during surgery. Previous studies indicate a wide range of infusion rates (0.05–1.0 μg/kg/min) when used in combination with other anesthetics [37], and individual responses to remifentanil vary based on factors such as patient gender, weight, and underlying conditions, necessitating skilled anesthesiologists for appropriate titration [38]. It is important to reduce the use of opioids during surgery to minimize these adverse events and improve prognosis, and the minimum use of remifentanil at the right time while maintaining sufficient analgesic effects in patients is required [39,40]. The results of this study will be considered clinically meaningful if the amount of remifentanil used in the acupuncture group decreases, even if the differences in blood pressure fluctuations or pulse rate changes are not significant between the two groups. Therefore, we set the amount of remifentanil used during the early stage of surgery, where blood pressure fluctuations were observed, as one of our secondary outcomes.

The greatest limitation of this study is that completeness can be suspected because the performers will not be blinded, as in many other acupuncture studies. To overcome this limitation, this study adopted a complementary method suggested in previous studies. By completely separating the assessments, an independent and blinded assessor will evaluate the outcomes.

The results of this study are expected to support the use of acupuncture therapy as another non-pharmacological intervention to contribute to surgery by maintaining blood pressure fluctuations within stable ranges during the early stages of surgery, which can be further validated in subsequent studies. This study is being timely updated in Clinicaltrials.gov. Once the documents are complete after the study ends, the study results would also be published.

## Supporting information

Supplement 1. protocol checklist

## Data Availability

All data produced in the present study are not available now. The publication is scheduled for December 2025. Data deposition is scheduled to occur.

https://datadryad.org/stash

## ETHICS AND DISSEMINATION

### Author Contributions

Conceptualization, Lee J.H. and Roh J; methodology, Han K.H. and Lee J.Y.; writing—original draft preparation, Lee J.H.; writing—review and editing, Yun B.S. and Na Y.J. Lee J.Y.; visualization, Lee J.Y.; supervision, Roh J; project administration, Na Y.J. Kim M.; funding acquisition, Lee J.Y. All authors have read and agreed to the published version of the manuscript.

### Trial Registration

This study was registered at Clinicaltrials.gov (registration identifier: NCT05720884) and CRiS (registration identifier: KCT0009149).

## Acknowledgments and Funding

This research is supported by a grant from the Korea Health Technology R&D Project through the Korea Health Industry Development Institute (KHIDI), funded by the Ministry of Health & Welfare, Republic of Korea (grant number: RS-2023-KH139140 (HF23C0097)).

## Institutional Review Board Statement

The study will be conducted in accordance with the Declaration of Helsinki, and approved by the Institutional Review Board of Cha Ilsan Medical Center (ICHA 2022-11-010, date of approval 2023-01-03).

## Informed Consent Statement

Informed consent will be obtained from all participants involved in the study. Written informed consent will be obtained from the patients to publish this paper if applicable.

## Publication and Data Deposition

The publication is scheduled for December 2025. Data deposition is scheduled to occur.

## Dissemination

In formulating the research question, we considered the patients’ reported discomfort and conducted a survey of hospital staff. Blood pressure fluctuation was not a primary symptom identified by the patients. A public hearing may be held after this pilot trial to engage the public opinion prior to initiating the main study.

## References

1. Jessen MK, Vallentin MF, Holmberg MJ; et al. Goal-directed haemodynamic therapy during general anaesthesia for noncardiac surgery: a systematic review and meta-analysis. Br J Anaesth 2022;128:416–33.

2. Roach J, Cha S. Monitoring during vascular surgery. Anesthesiol Clin 2022;40:645–55.

3. Mascha EJ, Yang D, Weiss S et al. Intraoperative Mean arterial Pressure Variability and 30-day Mortality in Patients Having Noncardiac Surgery. Anesthesiology 2015;123:79–91.

4. Juri T, Suehiro K, Kimura A et al. Impact of continuous non-invasive blood pressure monitoring on hemodynamic fluctuation during general anesthesia: a randomized controlled study. J Clin Monit Comput 2018;32:1005–13.

5. Taneja S, Jain A. Systematic review and meta-analysis comparing the efficacy of dexmedetomidine to midazolam as premedication and a sedative agent in pediatric patients undergoing dental procedures. Oral Maxillofac Surg 2023;27:547–57.

6. Park JW, Min BH, Park SJ et al. Midazolam premedication facilitates mask ventilation during induction of general anesthesia: a randomized clinical trial. Anesth Analg 2019;129:500–6.

7. Dwivedi P, Patel TK, Bajpai V et al. Efficacy and safety of intranasal ketamine compared with intranasal dexmedetomidine as a premedication before general anesthesia in pediatric patients: a systematic review and meta-analysis of randomized controlled trials. Can J Anaesth 2022;69:1405–18.

8. Kowark A, Rossaint R, Keszei AP; et al. Impact of preoperative midazolam on outcome of elderly patients (I-promote): study protocol for a multicentre randomised controlled trial. Trials 2019;20:430.

9. Shen F, Zhang Q, Xu Y et al. Effect of intranasal dexmedetomidine or midazolam for premedication on the occurrence of respiratory adverse events in children undergoing tonsillectomy and adenoidectomy: A randomized clinical trial. JAMA Netw Open 2022;5:e2225473.

10. Hahn DW, Lee CH, Chon KK. Korean adaptation of Spielberger’s STAI (KSTAI). Korean J Health Psychol 1996;1:1–14.

11. Lee K, Bae H, Kim D. Factor analysis of the Korean version of the State–Trait Anxiety Inventory in patients with anxiety disorders. Anxiety Mood 2008;4:104–10.

12. Kim MH, Cho YS, Uhm WS et al. Cross-cultural adaptation and validation of the Korean version of the EQ-5D in patients with rheumatic diseases. Qual Life Res 2005;14:1401–6.

13. Gregory A, Stapelfeldt WH, Khanna AK, et al. Intraoperative hypotension is associated with adverse clinical outcomes after noncardiac surgery. Anesth Analg 2021;132:1654–65.

14. Messina A, Robba C, Calabro L, Zambelli D, et al. Association between perioperative fluid administration and postoperative outcomes: a 20-year systematic review and a meta-analysis of randomized goal-directed trials in major visceral/noncardiac surgery. Crit Care 2021;25:43.

15. Halvorsen S, Mehilli J, Cassese S, et al. 2022 ESC Guidelines on cardiovascular assessment and management of patients undergoing non-cardiac surgery. Eur Heart J 2022;43(39):3826–924. doi: 10.1093/eurheartj/ehac270 [published Online First: 2022/08/27.

16. Beverly A, Kaye AD, Ljungqvist O et al. Essential Elements of Multimodal Analgesia in Enhanced Recovery After Surgery (ERAS) Guidelines. Anesthesiol Clin 2017;35:e115–43.

17. Park S, Lee HC, Jung CW; et al. Intraoperative arterial pressure variability and postoperative acute kidney injury. Clin J Am Soc Nephrol 2020;15:35–46.

18. Futier E, Lefrant JY, Guinot PG, et al. Effect of individualized vs standard blood pressure management strategies on postoperative organ dysfunction among high-risk patients undergoing major surgery: a ran-domized clinical trial. JAMA 2017;318:1346–57.

19. Yeh BY, Chao YL, Chen YS et al. Effect of acupuncture on capillary refill time in healthy adults: a clinical study. Microvasc Res 2021;135:104135.

20. Zhang J, Lyu T, Yang Y et al. Acupuncture at LR3 and KI3 shows a control effect on essential hypertension and targeted action on cerebral regions related to blood pressure regulation: a resting state functional magnetic resonance imaging study. Acupunct Med 2021;39:53–63.

21. Flachskampf FA, Gallasch J, Gefeller O et al. Randomized trial of acupuncture to lower blood pressure. Circulation 2007;115:3121–9.

22. Yin C, Seo B, Park HJ et al. Acupuncture, a promising adjunctive therapy for essential hypertension: a double-blind, randomized, controlled trial. Neurol Res 2007;29 Supplement 1:S98–SS103.

23. Huang F, Yao GX, Huang XL et al. Clinical observation on acupuncture for treatment of hypertension of phlegm-stasis blocking collateral type. Zhongguo Zhen Jiu 2007;27:403–6.

24. Chen J, Lee J, Wang ZR. Therapeutic effect on essential hypertension treated with combined therapy of acupuncture and medication. Zhongguo Zhen Jiu 2010;30:896–8.

25. Bäumler P, Zhang W, Stübinger T et al. Acupuncture-related adverse events: systematic review and meta analyses of prospective clinical studies. BMJ Open 2021;11:e045961.

26. Bijker JB, van Klei WA, Kappen TH et al. Incidence of intraoperative hypotension as a function of the chosen definition: literature definitions applied to a retrospective cohort using automated data collection. Anesthesiology 2007;107:213–20.

27. van Klei WA, van Waes JAR, Pasma W et al. Relationship between preoperative evaluation blood pressure and preinduction blood pressure: a cohort study in patients undergoing general anesthesia. Anesth Analg 2017;124:431–7.

28. Saugel B, Reese PC, Sessler DI et al. Automated ambulatory blood pressure measurements and intraoperative hypotension in patients having noncardiac surgery with general anesthesia: a prospective observational study. Anesthesiology 2019;131:74–83.

29. Bijker JB, van Klei WA, Vergouwe Y et al. Intraoperative hypotension and 1 year mortality after noncardiac surgery. Anesthesiology 2009;111:1217–26.

30. Min JY, Kim HI, Park SJ et al. Adequate interval for the monitoring of vital signs during endotracheal intubation. BMC Anesthesiol 2017;17:110.

31. Venkatraman R, Chitrambalam TG, Preethi A. Comparison of different carbon dioxide insufflation rates on hemodynamic changes in laparoscopic surgeries: A randomized controlled trial. Cureus 2023;15:e34071.

32. Yoon HK, Bae H, Yoo S et al. Relationships between common carotid artery blood flow and anesthesia, pneumoperitoneum, and head-down tilt position: a linear mixed-effect analysis. J Clin Monit Comput 2023;37:669–77.

33. Heyba M, Khalil A, Elkenany Y. Severe intraoperative bradycardia during laparoscopic cholecystectomy due to rapid peritoneal insufflation. Case Rep Anesthesiol 2020;2020:8828914.

34. Schumacher M, Fukuda K. Opioids. In: Gropper MA, Eriksson LI, Fleisher LA et al., Eds. Miller’s anesthesia. 9th ed.–741.e15. Amsterdam, NL: Elsevier Health Sciences 2019, 680.

35. Chen L, He W, Liu X et al. Application of opioid-free general anesthesia for gynecological laparoscopic surgery under ERAS protocol: a non-inferiority randomized controlled trial. BMC Anesthesiol 2023;23:34.

36. Jin Y, Mao Y, Chen D; et al. Thalamocortical circuits drive remifentanil-induced postoperative hyperalgesia. J Clin Invest 2022;132:e158742.

37. Eleveld DJ, Colin P, Absalom AR et al. Target-controlled-infusion models for remifentanil dosing consistent with approved recommendations. Br J Anaesth 2020;125:483–91.

38. Minto CF, Schnider TW, Egan TD; et al. Influence of age and gender on the pharmacokinetics and pharmacodynamics of remifentanil. I. Model development. Anesthesiology 1997;86:10–23.

39. Bugada D, Lorini LF, Lavand’homme P. Opioid free anesthesia: evidence for short and long-term outcome. Minerva Anestesiol 2021;87:230–7.

40. Beloeil H. Opioid-free anesthesia. Best Pract Res Clin Anaesthesiol 2019;33:353–60.

